# Virome of New York Presbyterian Hospital pediatric emergency

**DOI:** 10.1101/2020.03.18.20038166

**Authors:** Minhaz Ud-Dean, Ioan Filip, Marta Galanti, Ruthie Birger, Devon Comito, Gregory Freyer, Sadiat Ibrahim, Benjamin Lane, Chanel Ligon, Nelsa Matienzo, Haruka Morita, Atinuke Shittu, Eudosie Tagne, Peter Dayan, Jeffrey Shaman

## Abstract

**Background:** Viral infection of the respiratory tract is one of the major causes of hospital visits for young children. In this study, we report the occurrence and co-occurrence of different virus types and subtypes among the patients arriving at the pediatric emergency room of New York Presbyterian Hospital, a major urban hospital.

**Methods:** We collected nasal swabs from the patients and their accompanying persons. We also recorded the levels (None, Mild, High, and Severe) of their symptoms (Fever, Chill, Muscle Pain, Watery Eye, Runny Nose, Sneezing, Sore throat and Cough). The collected swabs were tested for the presence of common viruses infecting the respiratory tract.

**Results:** Human Rhinovirus was the most common virus among the patients, followed Influenza and Respiratory Syncytial Virus. Human Rhinovirus was most common in summer, autumn and spring. In contrast, influenza was more common in winter. Further, Influenza A virus was more likely to co-occur with Corona Virus 229E. In comparison, Influenza was less likely to co-occur with Human Rhinovirus. Moreover, Influenza, Parainfluenza and Corona virus were associated with more severe symptoms, while Human Rhinovirus was associated with less severe symptoms. In addition, we observed that Influenza and Respiratory Syncytial Virus were more likely to infect a patient when these viruses also infected the accompanying person. We also found that it was difficult to distinguish among viruses based on the symptoms. The inability to distinguish among different virus types and subtypes is explained by the fact that multiple viruses lead to similar symptoms.

**Conclusions:** The findings of this study provide a better understanding of respiratory viral infections in small children presenting at a pediatric emergency room in New York.

## Background

Respiratory Tract Infections (RTIs) are among the most common illnesses irrespective of age [1], and a major cause of child mortality [2, 3]. Viral RTI spread more readily among small children, whose lifetime exposure to these pathogens is more limited and whose immune systems may be less developed. Previous large-scale studies in both children and adults have demonstrated differences among viral RTI agents in terms of infection rates, seasonality and severity of symptoms [4-6]. Additionally, the prevalence of co-infection by multiple viruses has also been reported in certain settings [7]. However, the incidence correlation of viral infections as well as the asymptomatic infection rates in family members of infected children remains poorly understood.

In this study, nasal swabs and symptomology were collected from patients arriving at the pediatric emergency room (PedER) of New York Presbyterian Hospital (NYPH). Swabs were assayed for the presence of different viral types and subtypes. Here we report the number of patients infected by each virus, the interdependence of viral infections expressed as conditional infection rates i.e. whether infection by a certain virus makes a patient more likely to be infected by another virus, shared symptomatic and asymptomatic infection rates among accompanying family members, and the differences in symptoms among different viruses.

## Methods

In this cross-sectional study, participants were enrolled between 1 Aug 2016 and 4 Aug 2017 at the pediatric emergency department of NYPH, a single, urban pediatric emergency department with an annual volume of approximately 50,000 patients. Children below the age of 18 presenting with respiratory complaints (i.e. acute illness, asthma) and their accompanying adult (e.g. parents or other relatives) were provided a detailed study description and if interested, provided consent to participate. The consenting adult filled out a questionnaire for themselves and the child, and two nasopharyngeal samples were collected from each participant (accompanying adult and the child with complaints).

### Questionnaire

The questionnaire included information on ethnicity, general health (i.e. if they often feel sick), daily habits, travel history in the previous month and household structure. Nine respiratory illness-related symptoms (fever, chills, muscle pain, watery eyes, runny nose, sneezing, sore throat, cough, chest pain during the past 48 hours) were also recorded on a Likert scale (0=none, 1=mild, 2=moderate, 3=severe).

### Specimen collection and analysis

From each participant two nasopharyngeal samples were collected using minitip flocked swabs. Both the samples were stored in a single tube in 2ml DNA/RNA Shield (Zymo Research, Irvine, CA) at 4-25°C for up to 30 days. The samples were then split into two aliquots, and stored at -80°C. Total nucleic acid extraction was completed using the easyMAG NucliSENS (Biomerieux, Durham, NC). The eSensor XT-8 Respiratory Viral Panel (RVP; GenMark Diagnostics, Carlsbad, CA), a multiplex PCR assay, was used for virus detection according to the manufacturer’s instructions. The RVP separately detects influenza (Inf) A (any subtype: A/H1N1, A/H3N2, A/H1N1pdm2009), influenza B, Respiratory Syncytial Virus (RSV) A and B, parainfluenza (PIV) 1, 2, 3, and 4, human metapneumovirus (hMPV), human rhinovirus (HRV), adenovirus B/E and C, and coronavirus (CV) 229E, NL63, OC43, and HKU1. The eSensor system measures electrical current density in nA/mm^2^. Samples that produced current density ≥2 nA/mm^2^ for a particular virus were considered positive except for CV OC43. According to protocol of GenMark, the threshold for considering a sample positive for CV OC43 was 25 nA/mm^2^.

We used numpy and scipy statistics packages to calculate mean symptom scores, the coinfection rates (i.e. fraction of patients infected simultaneously by two viruses), conditional infection rates and statistical significances from the data. The mean symptom scores were calculated by taking the arithmetic mean of the symptom scores, and coinfection rates were calculated by dividing the number of coinfections by the total number of patients. The conditional infection rate of any virus A given infection by virus B was calculated by dividing the number of A and B coinfections by the number of infection by virus B, and finally we used Fisher’s exact test with Bonferroni correction to estimate the statistical significance of the conditional infection rates.

## Results

### Patients characteristics

340 patients (i.e. children below age 18 years presenting at the PedER) between the ages of 2 months and 18 years (mean ∼6 years) were enrolled between 1 Aug 2016 and 4 Aug 2017. The number of adults accompanying the patients was 324. The majority (62%) of patients were <5 years old. There were included 174 male, 165 female and 1 transgender patient. Number of patients varied per season: summer, 67 patients; autumn, 91 patients; winter, 89 patients; and spring, 92 patients. The number of patients shedding each virus is depicted in Table 1. HRV was the most common virus followed by influenza (all types) and respiratory RSV B.

**Table 1:** Number of patients shedding different virus.

| **Virus** | **Total** | **Summer** | **Autumn** | **Winter** | **Spring** |
| --- | --- | --- | --- | --- | --- |
| Inf | 47 | 0 | 4 | 29 | 14 |
| Inf A | 29 | 0 | 1 | 26 | 2 |
| Inf B | 18 | 0 | 3 | 3 | 12 |
| RSV A | 17 | 0 | 4 | 13 | 0 |
| RSV B | 18 | 0 | 6 | 10 | 1 |
| PIV 1 | 12 | 7 | 0 | 4 | 1 |
| PIV 2 | 10 | 1 | 7 | 2 | 0 |
| PIV 3 | 17 | 6 | 2 | 0 | 9 |
| PIV 4 | 3 | 3 | 0 | 0 | 0 |
| hMPV | 15 | 2 | 4 | 6 | 3 |
| HRV | 84 | 12 | 37 | 7 | 27 |
| Adeno B | 5 | 0 | 3 | 1 | 1 |
| Adeno C | 15 | 2 | 7 | 3 | 3 |
| CV 229E | 10 | 0 | 1 | 7 | 2 |
| CV NL63 | 1 | 0 | 0 | 0 | 1 |
| CV HKU1 | 2 | 0 | 0 | 0 | 2 |
| CV OC43 | 5 | 0 | 0 | 2 | 3 |

Well-known seasonal patterns were observed. For example, RSV A, RSV B and Inf A were most common in winter, whereas influenza B and PIV 3 were most common in spring. HRV occurred all year, but was more common in autumn and spring (Table 1**Error! Reference source not found**.). Note that due to the ambiguity in determining whether a person shedding a virus is infected or not, in this study we use the terms ‘shedding a virus’ and ‘infected by a virus’ interchangeably.

### Infection rates are dependent on viral interactions

The infection rates of different viruses were correlated with infection by other viruses (Table 2). For instance, 8% of all patients were infected by Inf A**Error! Reference source not found**.; however, 50 % of the patients infected by CV 229E were also infected by Inf A (i.e. the conditional infection rate of Inf A given CV 229E was significantly different from random chance, P<0.001 Bonferroni corrected Fisher’s exact test). The most striking feature of the conditional infection rates was the almost complete exclusion between Inf and HRV. HRV (24%) and Inf A (8%) were both very common infections among the subjects. Thus, one would expect the number of patients coinfected by both HRV and Inf A to be approximately 2% of the patients; however, the observed coinfection rate was close to zero (P<0.01, Bonferroni corrected Fisher’s exact test).

**Table 2:** Conditional infection rates: The count row indicates the number of observed cases for each virus in the columns. The numbers in the infection rates row are calculated by dividing the count by the number of patients. The rows with virus names in the leftmost column indicate the conditional infection rates of the virus in the column given infection by the virus in the row. Only viruses presenting at least 15 counts are shown in this table. Rates significantly different from expected are shown in bold.

|  | Inf | InfA | InfB | RSV A | RSV B | PIV 3 | hMPV | HRV | Adeno C |
| --- | --- | --- | --- | --- | --- | --- | --- | --- | --- |
| **Count** | **47** | **29** | **18** | **17** | **18** | **17** | **15** | **84** | **15** |
| **Infection rate** | **0.14** | **0.08** | **0.05** | **0.05** | **0.05** | **0.05** | **0.04** | **0.24** | **0.04** |
| **Inf** |  | 0.62 | 0.38 | 0.00 | 0.02 | 0.00 | 0.00 | 0.02 | 0.04 |
| **Inf A** |  |  | 0.00 | 0.00 | 0.03 | 0.00 | 0.00 | **0.00** | 0.03 |
| **Inf B** |  | 0.00 |  | 0.00 | 0.00 | 0.00 | 0.00 | 0.06 | 0.06 |
| **RSV A** | 0.00 | 0.00 | 0.00 |  | 0.00 | 0.06 | 0.00 | 0.06 | 0.06 |
| **RSV B** | 0.06 | 0.06 | 0.00 | 0.00 |  | 0.00 | 0.00 | 0.00 | 0.06 |
| **PIV 1** | 0.08 | 0.08 | 0.00 | 0.00 | 0.00 | 0.00 | 0.00 | 0.00 | 0.00 |
| **PIV 2** | 0.00 | 0.00 | 0.00 | 0.10 | 0.00 | 0.00 | 0.00 | 0.50 | 0.00 |
| **PIV 3** | 0.00 | 0.00 | 0.00 | 0.06 | 0.00 |  | 0.06 | 0.18 | 0.06 |
| **PIV 4** | 0.00 | 0.00 | 0.00 | 0.00 | 0.00 | 0.00 | 0.00 | 0.33 | 0.00 |
| **hMPV** | 0.00 | 0.00 | 0.00 | 0.00 | 0.00 | 0.07 |  | 0.07 | 0.00 |
| **HRV** | 0.01 | **0.00** | 0.01 | 0.01 | 0.00 | 0.04 | 0.01 |  | 0.05 |
| **Adeno B** | 0.00 | 0.00 | 0.00 | 0.20 | 0.00 | 0.00 | 0.00 | 0.20 | 0.40 |
| **Adeno C** | 0.13 | 0.07 | 0.07 | 0.07 | 0.07 | 0.07 | 0.00 | 0.27 |  |
| **CV 229E** | 0.50 | **0.50** | 0.00 | 0.00 | 0.00 | 0.00 | 0.00 | 0.10 | 0.00 |
| **CV NL63** | 0.00 | 0.00 | 0.00 | 0.00 | 0.00 | 0.00 | 0.00 | 0.00 | 0.00 |
| **CV HKU1** | 0.00 | 0.00 | 0.00 | 0.00 | 0.00 | 0.00 | 0.00 | 0.00 | 0.50 |

### Infection in the Family

Having an infected family member increased the chance of infection by the same virus up to ten times (Fig. 1). Compared to the infection rates among all patients (i.e. fraction of all patients shedding a virus), the infection rates among patients with infected family member(s) were significantly different (P<0.001) for Inf A, Inf B, RSV A, and RSV B as tested by Bonferroni corrected Fisher’s exact test.

**Fig 1.**
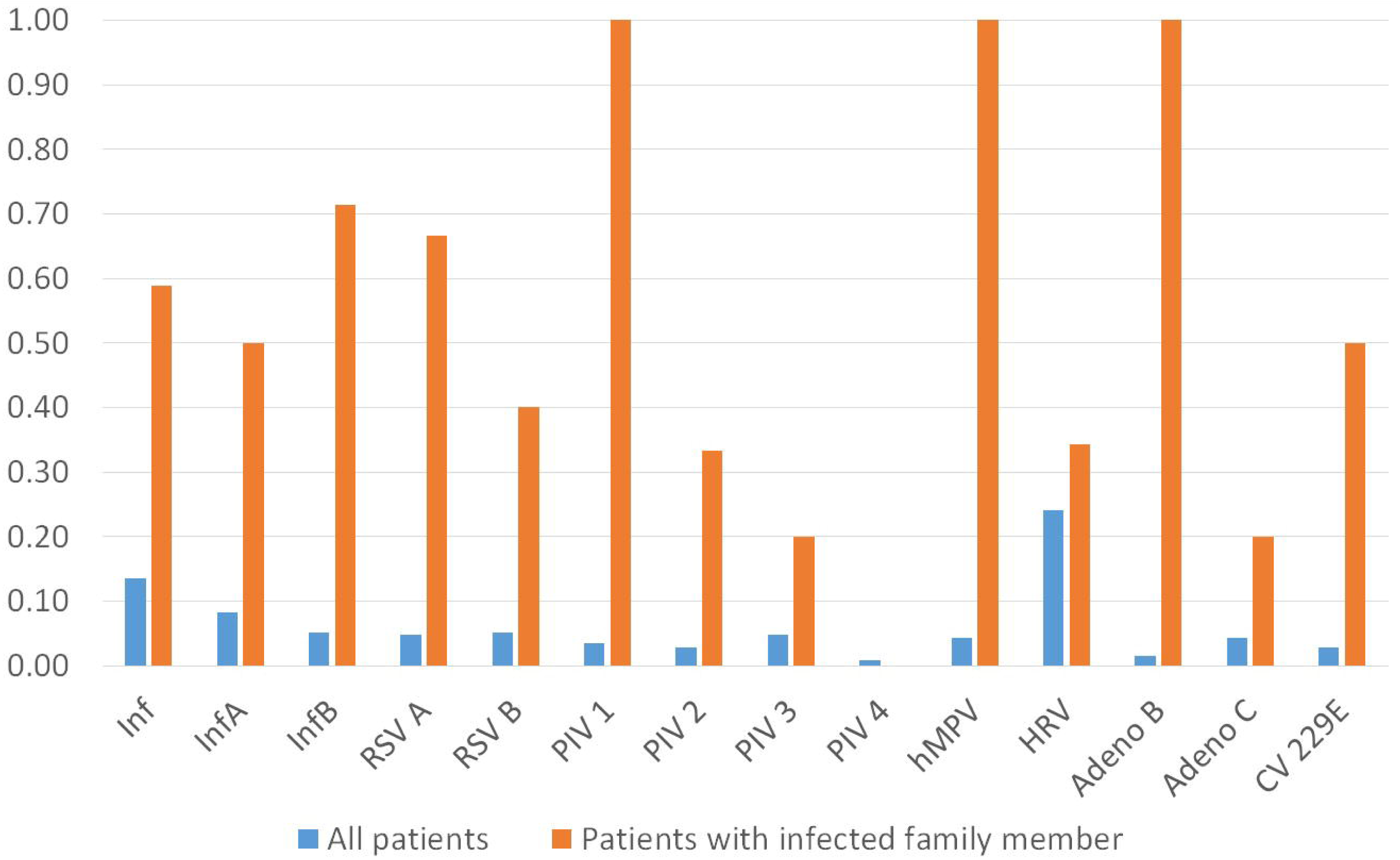
Likelihood a patient is infected by the same virus as an accompanying family member (orange) versus the overall pool of patients (blue).

As shown in table 3, some of the infected family members were asymptomatic. The rate of asymptomatic infection was highest for HRV followed by RSV and Inf.

**Table 3:** Asymptomatic infection rates of different viruses among the companions of the patients. The asymptomatic infection rate is calculated by dividing the number of asymptomatic infections by the number of companions. An asymptomatic infection is defined in 4 different ways. Definition 1: total symptoms score less than 5, definition 2: total symptoms score less than 10, definition 3: total symptoms score less than 15, and definition 4: none of the 9 recorded symptoms is severe (i.e. score 3).

|  | Definition 1 |  | Definition 2 |  | Definition 3 |  | Definition 4 |  |
| --- | --- | --- | --- | --- | --- | --- | --- | --- |
|  | Count | Rate | Count | Rate | Count | Rate | Count | Rate |
| **Inf** | 12 | 0.037037 | 14 | 0.04321 | 14 | 0.04321 | 7 | 0.021605 |
| **InfA** | 5 | 0.015432 | 7 | 0.021605 | 7 | 0.021605 | 2 | 0.006173 |
| **InfB** | 7 | 0.021605 | 7 | 0.021605 | 7 | 0.021605 | 5 | 0.015432 |
| **RSV A** | 7 | 0.021605 | 11 | 0.033951 | 12 | 0.037037 | 6 | 0.018519 |
| **RSV B** | 10 | 0.030864 | 10 | 0.030864 | 10 | 0.030864 | 10 | 0.030864 |
| **PIV 1** | 4 | 0.012346 | 4 | 0.012346 | 4 | 0.012346 | 4 | 0.012346 |
| **PIV 2** | 3 | 0.009259 | 3 | 0.009259 | 3 | 0.009259 | 3 | 0.009259 |
| **PIV 3** | 5 | 0.015432 | 5 | 0.015432 | 5 | 0.015432 | 3 | 0.009259 |
| **PIV 4** | 1 | 0.003086 | 1 | 0.003086 | 1 | 0.003086 | 1 | 0.003086 |
| **hMPV** | 2 | 0.006173 | 2 | 0.006173 | 2 | 0.006173 | 2 | 0.006173 |
| **HRV** | 31 | 0.095679 | 33 | 0.101852 | 34 | 0.104938 | 30 | 0.092593 |
| **Adeno B** | 2 | 0.006173 | 2 | 0.006173 | 2 | 0.006173 | 1 | 0.003086 |
| **Adeno C** | 3 | 0.009259 | 3 | 0.009259 | 4 | 0.012346 | 2 | 0.006173 |
| **CV 229E** | 1 | 0.003086 | 4 | 0.012346 | 4 | 0.012346 | 1 | 0.003086 |

### Knowledge of seasonal viruses improves symptomatic diagnosis

The symptoms experienced varied by virus and season. As shown in Fig. 2, influenza (both A and B) caused severe fever in autumn, but more moderate fever in winter and spring. In contrast, PIV 2 caused severe fever in summer but mild fever during winter and spring. HRV was associated with rather mild fever in all seasons. RSV A and RSV B produced more severe fever during autumn but milder fever during winter.

**Fig 2:**
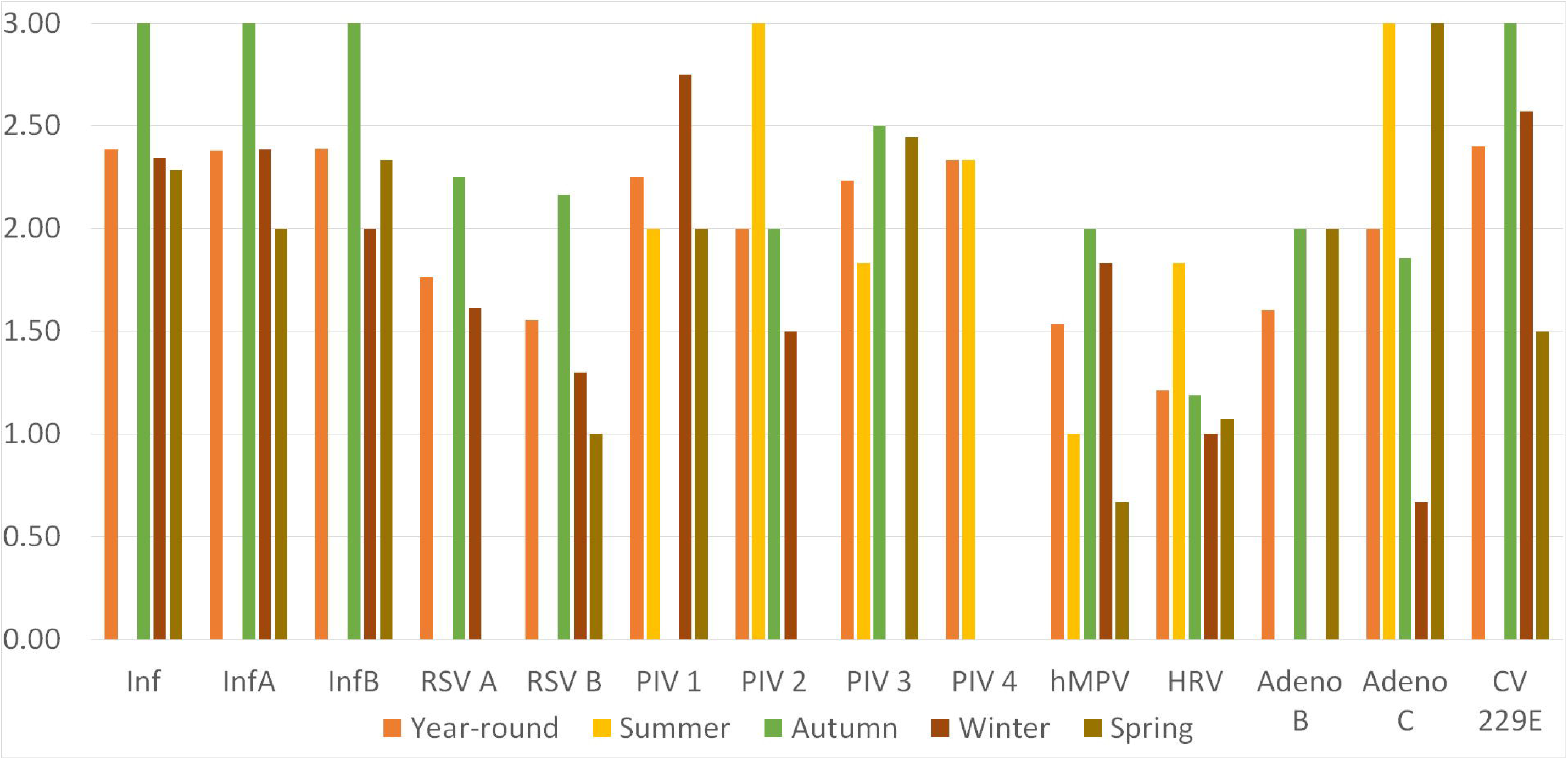
Average fever score associated with viral infections during different seasons as measured according to the self-reported scale varying from 0 to 3.

Diagnosing the causative agent of RTI from symptoms is challenging due to the similarity of the symptoms exhibited by different respiratory viruses. However, despite the similarity of symptoms, knowledge of background infection rates and severity of specific symptoms was useful for diagnosing viral infection type. As an example, we compared the likelihood of infection by Influenza (both A and B) and HRV. Among patients year-round, the infection rate of Inf was 14% and that of HRV was 24%, but among patients exhibiting moderate to severe muscle pain Inf was more likely (29%) than HRV (21%). In contrast, patients with moderate to severe chest pain had a 33% chance of HRV infection compared to an 8% chance of Inf. However, using models to predict of infection based on the symptoms taking into account overall infection rates for the past 4 weeks were ineffective in distinguishing different viruses. This indicated that accurate diagnosis of a specific respiratory infectious agent by symptoms is limited.

## Discussion

Our findings in this study of patients at the NYPH PedER are consistent with previous reports of seasonal viral infections and varying degrees of severity of symptoms based on the virus [4-6, 8]. Moreover, this study also provides data on the likelihood of infection by a given virus when patients exhibit different symptoms. Moreover, the most frequently reported viruses (HRV, Inf, RSV, ADV and PIV) from this study matches with those from Wang et al. [6]; however, the most prevalent virus in this study was HRV rather than RSV as reported by Wang et al. [6].

In this study, we found that influenza, parainfluenza and corona virus were associated with more severe symptoms. Among the symptoms fever, runny nose, cough and sneeze were frequent while chest pain, muscle pain and watery eye were less common. However, symptom severity varied seasonally for a given virus. Seasonality of different viruses among the subjects of this study followed the well-known seasonal patterns. For instance, in the overall yearly data, HRV was most common followed by Inf A and RSV. However, these viruses had very different seasonal prevalence. HRV was most common in summer, autumn and spring with influenza being most common in winter.

Our finding suggested that Inf B, RSV A, PIV 1, hMPV, and Adeno B were more likely to infect children when the accompanying family members were also infected. Additionally, more than 10% of the accompanying family members were infected by respiratory viruses despite showing no symptoms.

Examination of conditional infection rates indicates viral interaction. In particular influenza infection was more common among patients with CV 229E and appeared to be inhibited among patients with HRV. The exact reason behind this is not clear. The present study is limited by the number of patients. Further study of such viral facilitation and inhibition with larger sample sizes might clarify this interesting phenomenon.

## Conclusions

This study provides insights into occurrence, co-occurrence and asymptomatic infection rates of respiratory viruses in New York. The results indicated that HRV, RSV and Inf were the most common viruses in pediatric emergency setting. Despite both being common viruses, HRV and Inf coinfection was rare. Moreover, Inf B, RSV A, PIV 1, hMPV, and Adeno B were more likely to infect children when the accompanying family members were also infected. Additionally, around 10% of the accompanying adults were asymptomatically infected by respiratory viruses. These findings would facilitate future treatment and control of respiratory viral infections.

## Data Availability

The datasets generated and/or analyzed during the current study are not publicly available due to patient privacy but are available from the corresponding author on reasonable request.

## List of Abbreviation

CV: Corona Virus
hMPV: Human Metapneumovirus
HRV: Human Rhinovirus
Inf: Influenza
Inf: A Influenza A
Inf: B Influenza B
IRB: Institutional Review Board
NYPH: New York Presbyterian Hospital
PedER: Pediatric Emergency Room
PIV: Parainfluenza Virus
RSV: Respiratory Syncytial Virus
RTI: Respiratory Tract Infections
RVP: Respiratory Viral Panel

## Declarations

### Ethics approval and consent to participate

The study was approved by the institutional review board (IRB) of Columbia University. The IRB approval number is AAAQ4358.

### Consent for publication

Not Applicable.

## Authors’ contributions

JLS, GF and PD designed the study. JLS, HM, SI and PD oversaw collection of samples and questionnaires. DC, SI, BL, CL, NM, AS and ET analyzed the samples. MU and IF analyzed data. MU, JLS, PD, IF, MG, RB, HM and DC wrote the paper.

## Acknowledgements

Not Applicable.

## References

1. Monto AS: Epidemiology of viral respiratory infections. The American journal of medicine 2002, 112:4–12.

2. Hardelid P, Dattani N, Cortina-Borja M, Gilbert R: Contribution of respiratory tract infections to child deaths: a data linkage study. BMC Public Health 2014, 14:1191.

3. Rudan I, Chan KY, Zhang JS, Theodoratou E, Feng XL, Salomon JA, Lawn JE, Cousens S, Black RE, Guo Y: Causes of deaths in children younger than 5 years in China in 2008. The Lancet 2010, 375:1083–1089.

4. He Y, Lin GY, Wang Q, Cai XY, Zhang YH, Lin CX, Lu CD, Lu XD: A 3-year prospective study of the epidemiology of acute respiratory viral infections in hospitalized children in Shenzhen, China. Influenza and other respiratory viruses 2014, 8:443–451.

5. Wang H, Zheng Y, Deng J, Chen X, Liu P, Li X: Molecular epidemiology of respiratory adenovirus detection in hospitalized children in Shenzhen, China. International journal of clinical and experimental medicine 2015, 8:15011.

6. Wang H, Zheng Y, Deng J, Wang W, Liu P, Yang F, Jiang H: Prevalence of respiratory viruses among children hospitalized from respiratory infections in Shenzhen, China. Virology journal 2016, 13:39.

7. Calvo C, GarcÍa-GarcÍa ML, Pozo F, Paula G, Molinero M, Calderón A, González-Esguevillas M, Casas I: Respiratory syncytial virus coinfections with rhinovirus and human bocavirus in hospitalized children. Medicine 2015, 94.

8. Wang W, Cavailler P, Ren P, Zhang J, Dong W, Yan H, Mardy S, Cailhol J, Buchy P, Sheng J: Molecular monitoring of causative viruses in child acute respiratory infection in endemo-epidemic situations in Shanghai. Journal of Clinical Virology 2010, 49:211–218.

